# Machine learning assisted DSC-MRI radiomics as a tool for glioma classification by grade and mutation status

**DOI:** 10.1101/19007898

**Authors:** Carole H. Sudre, Jasmina Panovska-Griffiths, Eser Sanverdi, Sebastian Brandner, Vasileios K. Katsaros, George Stanjalis, Francesca B. Pizzini, Claudio Ghimenton, Katarina Surlan-Popovic, Jernej Avsenik, Maria Vittoria Spampinato, Mario Nigro, Arindam R. Chatterjee, Arnaud Attye, Sylvie Grand, Alexandre Krainik, Nicoletta Anzalone, Gian Marco Conte, Valeria Romeo, Lorenzo Ugga, Andrea Elefante, Elisa Francesca Ciceri, Elia Guadagno, Eftychia Kapsalaki, Diana Roettger, Javier Gonzalez, Timothé Boutelier, M. Jorge Cardoso, Sotirios Bisdas

## Abstract

**Background:** Machine learning assisted MRI radiomics, which combines MRI techniques with machine learning methodology, is rapidly gaining attention as a promising method for staging of brain gliomas. This study assesses the diagnostic value of such framework applied to dynamic susceptibility contrast (DSC)-MRI in classifying treatment-naïve gliomas from a multi-center patient pool into WHO grades II-IV and across their isocitrate dehydrogenase (IDH) mutation status.

**Methods:** 333 patients from 6 tertiary centres, diagnosed histologically and molecularly with primary gliomas (IDH-mutant=151 or IDH-wildtype=182) were retrospectively identified. Raw DSC-MRI data was post-processed for normalised leakage-corrected relative cerebral blood volume (rCBV) maps. Shape, intensity distribution (histogram) and rotational invariant Haralick texture features over the tumour mask were extracted. Differences in extracted features between IDH-wildtype and IDH-mutant gliomas and across three glioma grades were tested using the Wilcoxon two-sample test. A random forest algorithm was employed (2-fold cross-validation, 250 repeats) to predict grades or mutation status using the extracted features.

**Results:** Features from all types (shape, distribution, texture) showed significant differences across mutation status. WHO grade II-III differentiation was mostly driven by shape features while texture and intensity feature were more relevant for the III-IV separation. Increased number of features became significant when differentiating grades further apart from one another. Gliomas were correctly stratified by IDH mutation status in 71% of the cases and by grade in 53% of the cases. In addition, 87% of the gliomas grades predicted with an error distance up to 1.

**Conclusion:** Despite large heterogeneity in the multi-center dataset, machine learning assisted DSC-MRI radiomics hold potential to address the inherent variability and presents a promising approach for non-invasive glioma molecular subtyping and grading.

**Key points:**

- On highly heterogenous, multi-centre data, machine learning on DSC-MRI features can correctly predict glioma IDH subtyping in 71% of cases and glioma grade II-IV in 53% of the cases (87% <1 grade difference)
- Shape features distinguish best grade II from grade III gliomas.
- Texture and distribution features distinguish best grade III from grade IV tumours.

**Importance of study:** This work illustrates the diagnostic value of combining machine learning and dynamic susceptibility contrast-enhanced MRI (DSC-MRI) radiomics in classifying gliomas into WHO grades II-IV as well as across their isocitrate dehydrogenase (IDH) mutation status. Despite the data heterogeneity inherent to the multi-centre design of the studied cohort (333 subjects, 6 centres) that greatly increases the theoretical challenges of machine learning frameworks, good classification performance (accuracy of 53% across grades (87% <1 grade difference) and 71% across mutation status) was obtained. Therefore, our results provide a proof-of-concept for this emerging precision medicine field that has good generalisability and scalability properties. Introspection on the classification errors highlighted mostly borderline cases and helped underline the challenges of a categorical classification in a pathological continuum.

With its strong generalisability property, its ability to further incorporate participating centres and its possible use to identify borderline cases, the proposed machine learning framework has the potential to contribute to the clinical translation of machine-learning assisted diagnostic tools in neuro-oncology.

## Introduction

The most recent grading system of gliomas has integrated the mutation status of key encoding genes prompting a ground-breaking change in the context of glioma diagnosis and treatment ^1,2^ with further molecular stratification achieved by determining CDKN2A/B status^3^. Tumour anatomical features and gadolinium enhancement properties are not sufficient for this task and are extremely variable across different types and grades of gliomas with low specificity rates^4,5^. To overcome this limitation, advanced imaging techniques such as diffusion weighted imaging (DWI), MR spectroscopy, and dynamic susceptibility contrast (DSC)-MRI (also known as perfusion-weighted MRI) have been tested. Perfusion-weighted MRI provides biomarkers tightly linked to the tumour vascularity and thus indices of the biochemical and genetic substrates related to neo-angiogenesis^6,7^. ‘Gain-of-function’ isocitrate dehydrogenase (IDH) mutations result in the accumulation of the “oncometabolite” 2-hydroxy glutarate (HG) and cause an increase in the activity of the hypoxia-inducible factor-1, which has a pivotal role in the energy metabolism, angiogenesis and apoptosis^8^. Therefore, perfusion MRI biomarkers carry the potential to act as diagnostic surrogates of IDH mutation status and further for tumour grading.

DSC-MRI is often favoured, compared to other MRI-based perfusion techniques, for its higher temporal resolution and sensitivity for detecting abnormal angiogenesis resulting in high accuracy rates for baseline tumour grading and tumour surveillance^7^. To date, these encouraging findings have been mostly obtained in single-institution settings utilising different DSC-MRI acquisition techniques and post-processing software packages for quantitative analysis. This can result in significant variation^7^ and, hence, limit the diagnostic value of the technique impeding its large-scale application for non-invasive tumour phenotyping^9–11^. Recently, a consensus paper for DSC-MRI acquisition guidelines aimed to act as a blueprint and provide a framework for achieving routine success with this technique^12^.

The vast majority of the published DSC-MRI studies have focused on establishing the value of the tumour region of interest rather than encompassing the whole tumour. They have made use of histogram analyses, which nonetheless reflect numeric values of the voxels and only help to calculate first-order statistics. By contrast to first-order statistics, second- and higher-order statistical analyses, which allow measures of not only local voxel-wise values but also incorporate neighbouring information, have been suggested as more reliable and elaborative methods for characterisation of the region-of-interest^13^. Texture analysis, as a contextual quantification method, has already embarked in the medical imaging literature as a method that can detect tissue heterogeneity and complexity^13,14^. Radiomics, applied in the clinical context of glioblastoma, have generally been performed on anatomical sequences and applied to distinguish between GBM subtypes^15^, for prediction of survival rates^16^ and prognosis^17^, prediction of response to treatment^18^, as well as risk stratification^19^. Deep-learning based techniques have recently received a sustained attention due to their high performance in such classification tasks^20,21^. Nonetheless, there is limited biological insight in their decision process making failure cases difficult to interpret. An alternative is to use the classical feature-based machine learning methods as diagnostic tool and to evaluate the relevance of acquisition sequences. In such a framework, interaction between clinical interpretation and machine learning is essential. Indeed, an iterative loop between clinical interpretation and machine learning analysis based on advanced features can help to define most distinguishing images properties and exclude non-relevant ones. Such process would result in the definition of disease specific imaging signatures.

While the high heterogeneity among DSC-MRI acquisition protocols may contribute to lowering the diagnostic value of any single first-order statistic, this work sought to examine the role of higher-order analysis and machine-learning assisted radiomics in mitigating these limitations and augment the diagnostic accuracy of DSC-MRI for tumour staging with emphasis on IDH molecular subtyping. We hypothesise different combination of features are to be used according to the distinctions to be looked after encompassing either tissue heterogeneity between high grade GBM or measures of dissemination (using shape assessment) at lower grades. Compared to the work presented by Lu et al, where a publicly available database was used focusing solely on structural imaging, this work focuses for first time on the use of DSC-MRI acquisition in a multicentre setting. The heterogeneity inherent to the multi-centre quality of the data, makes the task of tumour and molecular phenotyping even more challenging. If successful, the proposed platform is readily expandable to include more participating institutions or utilising different machine learning approaches – all aiming to increase the diagnostic capacity and power to determine the most appropriate parameters for conducting DSC-MRI experiments. With this in mind, this work has potential to become a first step for a widespread application of machine-learning assisted diagnostic aid in neuro-oncology.

## Materials and methods

### Study design and ethics

This multi-center study was based on retrospectively identified or prospectively acquired neuro-oncology MRI data collection in the participating centres, according to the local institutional review boards guidelines and the respective local ethical committees’ approvals. Data sharing agreements between the collaborators and the central reading and post-processing site were also in place.

### Subjects

DSC-MRI data of 359 patients from 6 university hospitals with histopathologically and molecularly confirmed primary, treatment naïve gliomas (WHO grades II-IV) between January 2010 and May 2018 were consecutively collected and screened for this study. The inclusion criteria were as follows: i) existence of preoperative DSC-MRI examination raw data suitable for post-processing and ii) availability of histopathological and molecular testing least for IDH1 and IDH2 mutations^2^. Images with non-correctable motion artefacts (n= 8) and corrupt source data (n= 18) were excluded. Finally, 333 patients were included in the analysis.

### Histopathology

All gliomas in the cohort were diagnosed according to the WHO 2016 classification scheme, complemented with molecular profiling for IDH1 and IDH2 mutations^2,23–25^. The assessment of morphological features and immunostaining in the tumour samples was performed by neuropathologists in each institution. All IDH1R132H immune-negative gliomas were investigated by Sanger sequencing to identify out common tumour driver mutations of IDH1, IDH2, histone H3F3A, TERT promoter and BRAF genes^26,27^. 1q/19p codeletion status, EGFR amplification and 10q loss were determined by using a qPCR-based copy number assay in a subset of the participating institutions.

### MR Imaging

The eligible patients were examined on either 1.5T MR scanners (n=186, 56%) (Siemens SymphonyVision, Siemens Avanto, Siemens Healthineers, Erlangen, Germany; GE Discovery MR450, GE Healthcare, Chicago, USA; and Philips Achieva, Philips Medical Systems, Eindhoven, The Netherlands) or 3T MR units (n= 147, 44%) (Siemens Skyra, Siemens Allegra, Siemens TrioTim, Siemens Biograph_mMR, Siemens Healthineers, Erlangen, Germany; Philips Ingenia, Philips Medical Systems, Eindhoven, The Netherlands, 3T, Signa HDx, GE Healthcare, Waukesha, WI, USA).

### Tumour delineation and parametric modelling

Tumour segmentation was performed manually by the same radiologist, who was blind to the immunohistopathological results, on axial T2 (n=167, 49.7%) or axial FLAIR (n=169, 51.3%) series using ITK-SNAP Version 3.6.0^28^ excluding large cystic or necrotic tumour components and large vessels in line with previous relevant literature. When available, 3D T2/FLAIR series were preferred to their 2D counterparts.

Relative cerebral blood volume (rCBV) maps were generated from the DSC –MRI raw data using dedicated commercially available postprocessing software (Olea Sphere, Version 3; Olea Medical Solutions, La Ciotat, France). Perfusion source images motion correction, temporal and spatial filtering was applied in all cases. Subsequently, the software automatically defined the arterial input function (AIF) for each individual based on all voxels throughout the time series using global clustering method as it was described by Mouridsen et al^29^. Instead of the standard deconvolutional mathematical algorithms, a fully adaptive Bayesian scheme, which has been shown to be superior to the widely used classical approaches^30,31^, was chosen to derive the perfusion indices aiming at the most possibly accurate contrast agent leakage correction.

### Spatial and intensity normalisation

The customised pipeline for the post-processing of the tumour segmentations and perfusion (rCBV) images is presented in Figure 1a. The data was prepared for advanced analysis by isotropic resampling of the T1-weighted images followed by registration, normalisation of the parametric rCBV maps, and biomarker extraction through distribution and texture features. Isotropic resampling to 1mm^3^ voxels was facilitated using the NiftyReg open source software (https://sourceforge.net/projects/niftyreg) to allow for the computation of rotational invariant textural features. The T2-weighted/FLAIR images and rCBV maps were rigidly registered to the T1 isotropic image using NiftyReg^32^ and trilinear interpolation. The resulting co-registered maps were visually inspected to evaluate the success of the registration step. At the third stage, automatic normalisation of the parametric maps was performed in order to mitigate inter-subject variability and therefore, enable comparison of statistical and textural features across subjects. At this step, the basal ganglia of both hemispheres were initially segmented using the Geodesic Information Flows framework (GIF)^33^, a label fusion framework allowing for a local weighting of the information from a labelled database according to measures of local morphological similarities. The basal ganglia in the tumour-free hemisphere was used as a reference area for normalisation of the rCBV maps instead of healthy white matter in order to mitigate the risk of variability across the subjects due to white matter degeneration and vasculopathy. For tumours extending in the contralateral hemisphere, any tumour tissue was first removed from the reference region. Consequently, z-score maps over the tumour mask were derived with respect to the intensity distribution over the normalisation area.

**Figure 1:**
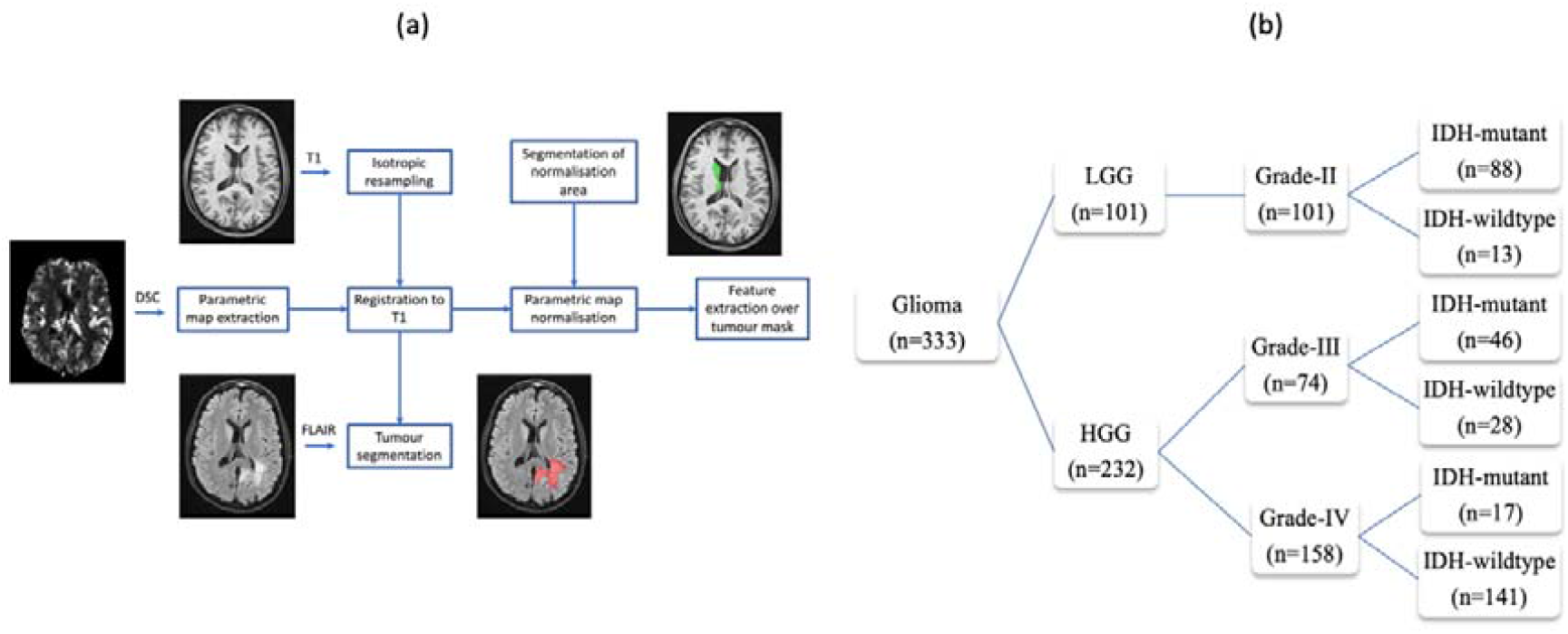
(a) Graphical illustration of the customised pipeline shows the cascade of processing starting from tumour segmentation on FLAIR series and ending in feature extraction over tumour mask. (b) Decision tree illustrating the different cohorts of gliomas from the multicentre dataset stratified per WHO grades II, III and IV and also per IDH mutation status.

### Feature extraction

Comprehensive biomarker extraction was performed using the normalised parametric maps. To capture tumour shape, four shape features (volume, surface/volume ratio, non-compactness) were extracted for each tumour mass. For distribution mapping, thirteen histogram features (mean, skewness, kurtosis, standard deviation, minimum, maximum, 1^st^, 5^th^, 25^th^, 50^th^ (median), 75^th^, 95^th^ and 99^th^ percentiles) were extracted through all voxels in each segmented tumour. Finally, the twelve Haralick features and including angular secondary moment (ASM), contrast, correlation, sum square, sum average, inverse difference moment (IDM), sum entropy, entropy, difference variance, sum variance, difference entropy, IMC1) for each tumour were also extracted^34^.

### Statistical analysis

All extracted intensity based features were corrected for the influence of MR acquisition parameters on the extracted features was scrutinised by scanner manufacturer, magnetic field strength (1.5T and 3T), TR (≤1499 ms and ≥1500 ms), TE(25-44 ms and 45-55 ms), FA (90° and <90°)slice thickness (<5 mm and ≥5 mm), and matrix size (matrix size <128×128 and ≥128×128) and in plane resolution (<=1, >1 and <=2, >=2). Following correction for these covariates, two-sample Wilcoxon test was used to report the differences in all extracted features between IDH mutation status and across pairs of grades. Results were considered to be significant if p-value ≤ 0.05 and all statistical analysis was carried out using Stata v14 and python 3.7. Effect size difference was calculated using Cliff’s Delta.

### Supervised learning from extracted features

All extracted features and acquisition parameters were used as input to a random forest^35^ to classify gliomas according to their IDH mutation status (IDH-mutant vs IDH-wildtype as a 2-class classifier) or according to their grade II, III and IV, as a 3-class problem. For each task (IDH mutation or grade assessment) training was performed in a stratified two-fold cross-validation setting with 250 repetitions to assess the value of rCBV-extracted features. Hyperparameters were optimised in a cross-validation setting leading to two different settings for the mutation task (200 trees, maximum depth of 10, minimum samples per leaf of 4) and the grade separation task (800 trees, minimum samples per leaf=4, maximum depth of 50). Error, computed as the difference between predicted and true classification was averaged over the 250 iterations and t-test over the error distribution were performed to assess the differences observed across acquisition parameters. Confusion matrices using the average classification were constructed to summarise the classification performance of the machine learning algorithm, from which the overall accuracy of classifying different gliomas cohorts, as well as overall sensitivity and specificity rates were calculated.

### Error introspection

In order to have more insight into the cases of erroneous classification, features were compared for each category of misclassification (wild type classified as mutant, mutant as wild type, grade II as grade III etc) and their rightly classified counterpart to investigate if there was a consistency in the features leading to the misclassification and whether or not it was consisted demographic findings between said classes. Wilcoxon 2 samples tests were performed on the features corrected for acquisition parameters, age and gender as per the demographic analysis.

## Results

### Demographic findings

Overall, data included 333 glioma patients comprising 198 males and 135 females with a mean age 48.9 years (age range 20–81 years). Of these, 101 were classified as grade II, 74 were grade III and 158 were grade IV while 151 were IDH-mutant and 182 were IDH-wildtype gliomas (Figure 1b). Details on the demographics of the patient population, tumour location, histological grades and the molecular mutation status are contained in table S1 of Appendix A.

Effect size of acquisition parameters are presented with the value of the Cliff Delta in supplementary table S2. Shape, histogram and texture features for comparing gliomas by mutation status and across three different grades are shown in Table 1, with Cliff’s Delta effects sizes presented pictorially in Figures 2.

**Table 1:**
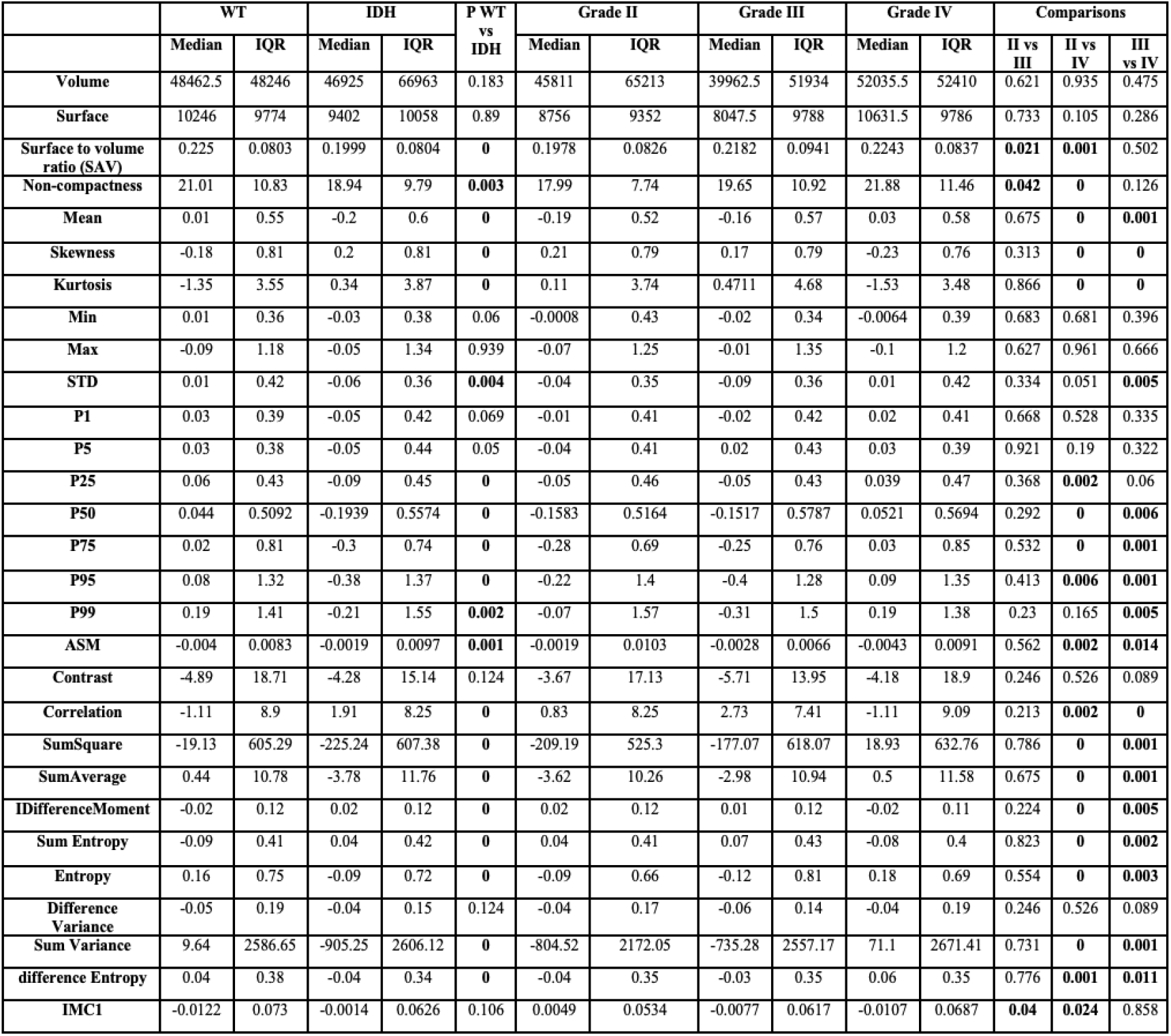
Median, IQR and p-value of features corrected for all acquisition parameters, age and gender. P-values of the Mann Whitney 2 samples test across IDH status and WHO grades II, III and IV are reported.

**Figure 2:**
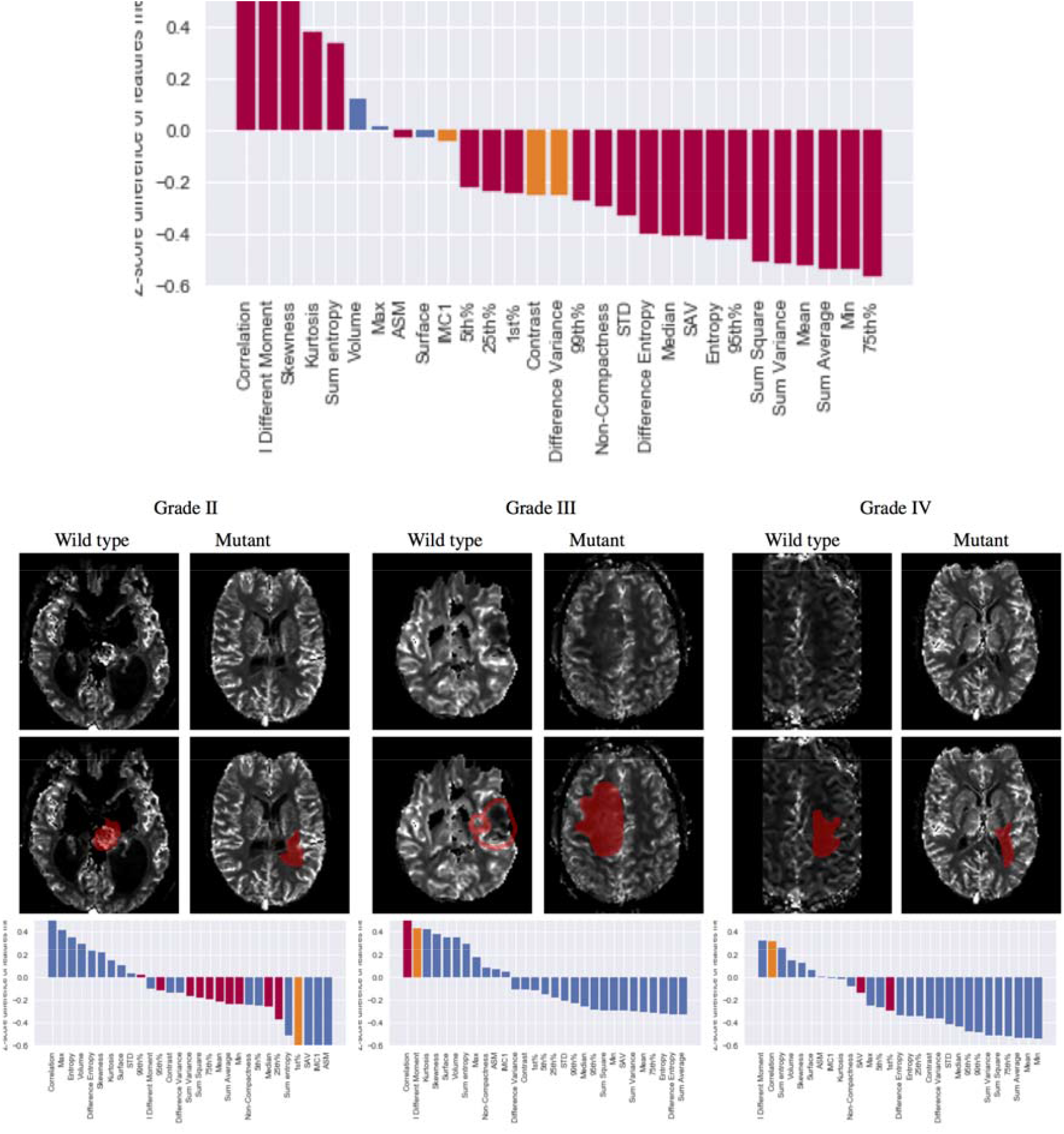
Comparisons of IDH wild type and IDH mutant gliomas within WHO grades II, III and IV gliomas. Typical examples of gliomas categories are shown for illustrator purposes on the top panel. Bar charts on the bottom panel present the mean z-score transformed difference between groups for each feature after correction for acquisition parameters for the non-shape features. Bars encoded in red represent features presenting significant statistical significance between groups, whereas bars in blue are those not statistically significant (significance threshold p=0.05).

**Figure 3:**
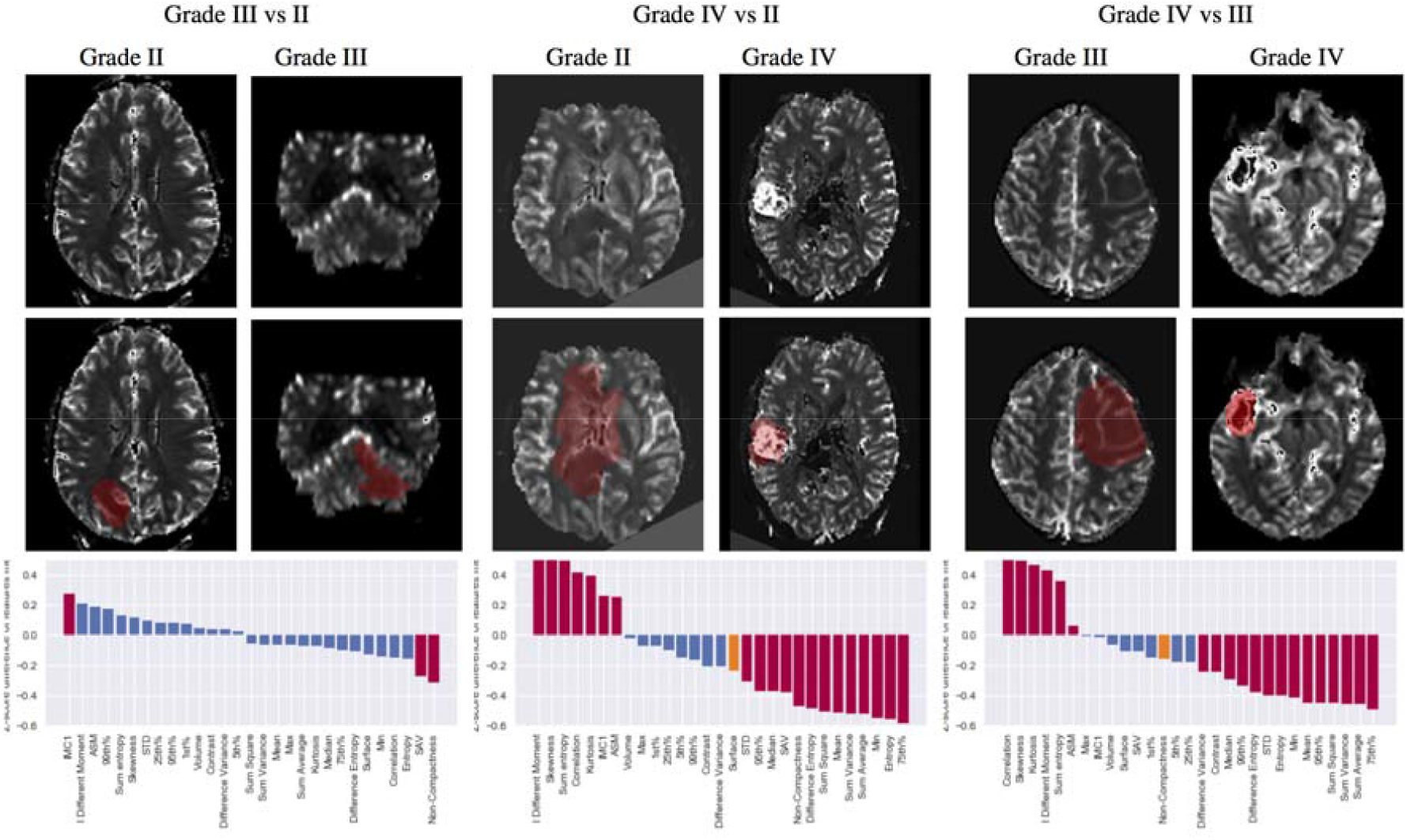
Comparisons between different WHO grade gliomas grades across shape, histogram and texture features. Typical examples of gliomas grades are shown for illustrator purposes on the middle panel. Bar charts on the top and bottom panel present the mean z-score transformed difference across all groups and between groups for each feature after correction for acquisition parameters for the non-shape features. Bars encoded in red represent features presenting significant statistical significance between groups as per Wilcoxon non parametric testing, whereas bars in blue are those not statistically significant (significance threshold p=0.05).

### Glioma stratification for IDH mutation status

When comparing gliomas by mutation type, 20 out of the 29 extracted features were significantly different between IDH-wildtype and IDH-mutants (column 6 on Table 1 and Figure 4A). Tumour surface to volume ratio (SAV) and measure of non-compactness were significantly lower in IDH-mutants compared to IDH-wildtype, as were six rCBV histogram features, including mean, standard deviation, 25^th^,50^th^,75^th^, 95^th^ and 99^th^ percentiles. Among the texture features, correlation and sum entropy were significantly higher in IDH-mutant tumours than in the IDH-wildtype (p<0.0001), while sum square, sum average, sum variance, entropy and difference variance features were significantly lower (p≤0.0001) in IDH-mutant gliomas compared to IDH wildtype gliomas.

**Figure 4:**
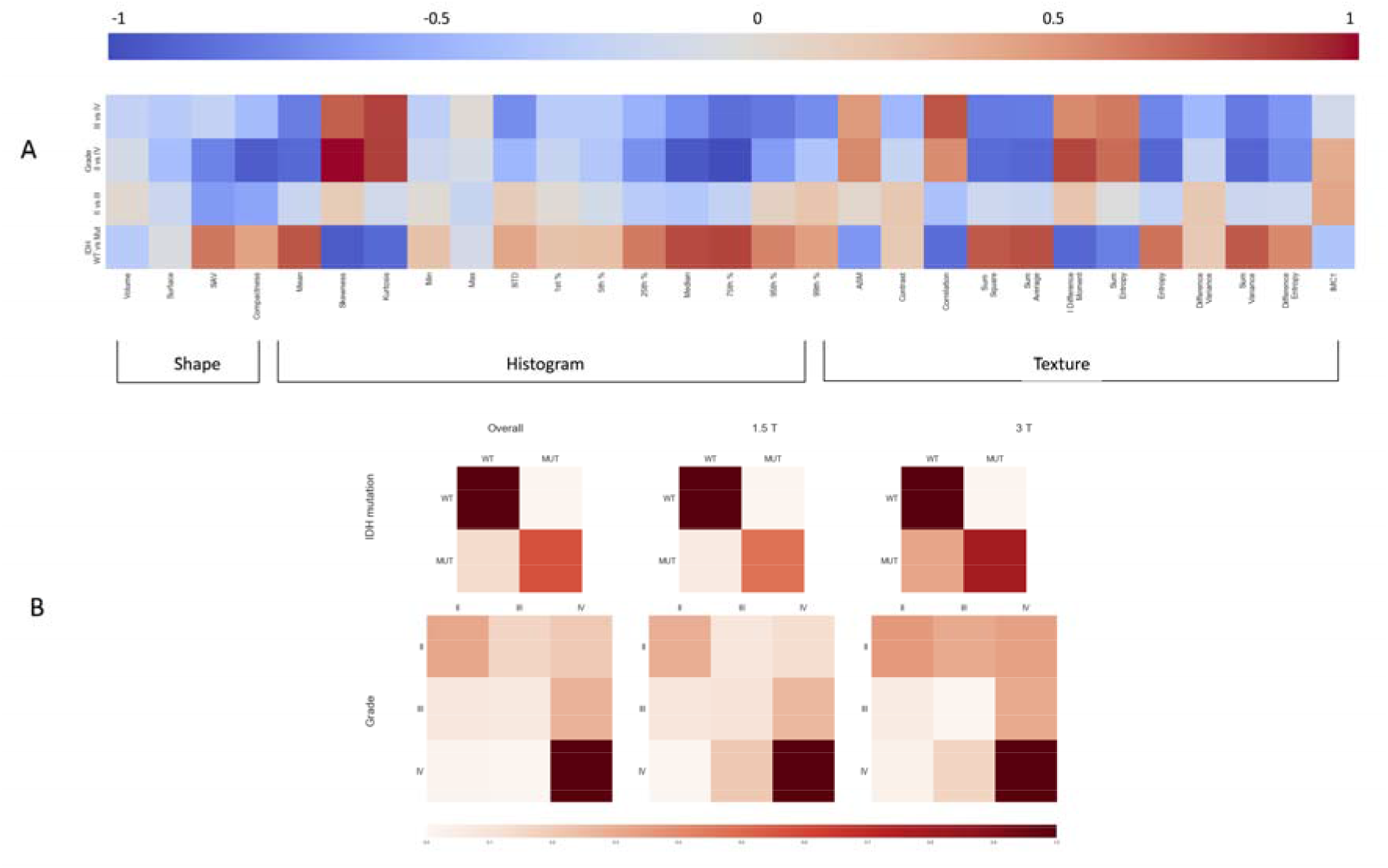
A: Pictorial representation of Cliff’s delta values when comparing features of shape, histogram and textures across mutation status and grades. Exact Cliff delta values are given in Table S1. Positive values are shown in darkening red while negative ones are in darkening blue. B: Outcomes of the confusion (or error) matrix aiding visualisation of the performance of our machine-learning algorithm. The first row shows the confusion matrices for the classification of gliomas by IDH mutation status. The second row shows the confusion matrices for the classification across the three WHO grades II, III and IV. In both classification scenarios we show three cases: using all data and separately using only data obtained from scanners with 1.5T or 3T magnetic field. Within each matrix, the matrix row represent the instances in the actual ground truth class while each column represents the instances in the predicted class. Darkening red correspond to higher percentages of the overall population to be classified. The exact values of the confusion matrices are given in supplementary table S4 of the Appendix.

### Glioma stratification for WHO-grade

When comparing gliomas grade III vs grade II, the surface to volume ratio shape feature, was significantly (p=0.021) higher in grade III gliomas compared to grade II gliomas as was the non-compactness feature (p=0.042) (Table 1, column 13). The IMC1 measure was also found to be lower in grade III compared to grade II (p=0.040)

The shape features did not differ between grades III and IV. Among the histogram features, 8 out of the 13 features were different with 5 (mean, STD, 50^th^, 75^th^, 95^th^ and 99tth percentile) higher and skewness and kurtosis lower in grade IV compared to grade III gliomas. Finally, among the texture features 9 out of 12 features were different. Notably texture features related to appearance heterogeneity such as the entropy, sum square and sum variance were larger in grade IV than III, while measures of correlation and inverse difference momentum were lower.

Comparing gliomas grades IV with II, significant differences could be observed in most of the extracted features (23 out of 29). Of the shape features, SAV and non-compactness were higher, with non-compactness of greater degree, in gliomas grade IV compared to grade II. Five histogram features (mean, 25^th^, 50^th^, 75^th^,95^th^ percentile) were higher while skewness and kurtosis were lower in grade IV compared to grade II gliomas. Finally, among the texture features, 10 out of 12 features, with only the contrast and the difference variance not presenting any statistically significant difference. Overall, comparisons between grade II and IV combined the differences observed between grade II and III and grade III and IV.

### Classification results

The confusion matrices describing the accuracy of the random forest algorithm are contained in the supplementary table S5 and are depicted in Figure 4B. The random forest algorithm correctly classified gliomas by IDH mutation status in 71% of the cases. This was slightly higher when we considered data from 1.5T scanner only (77% accuracy) while the accuracy dropped to 66% when we considered data from 3T scanner only. Stratifying gliomas by IDH mutation status had an overall specificity of 77% and specificity of 65%. When magnetic field strength was taken into consideration, at 1.5T the specificity rate increased up to 83% with a sensitivity at 70% while the specificity was of 72% and the sensitivity of 60% at 3T. Overall, we obtained significant (p=0.008) lower error rates of stratification at 1.5T (0.25) in comparison to 3T (0.35) (Table S3 and confusion matrices). Resolution appeared also to be associated with error, higher resolution leading to lower error rates (0.196 vs 0.320 p=0.024 for <=1mm vs between 1 and 2 mm) (see Table 3 Supplementary Table S2).

When classifying the gliomas by grade, 53% of included gliomas were classified correctly and 87% of the cases received grade classification with a distance less or equal to 1. The accuracy of classifying gliomas by grade was higher using perfusion data obtained at 1.5T magnetic field rather than at 3T: 59% and 46% respectively, p=0.024, with grade distance of less than 1 in 91% and 83% of the cases, respectively. Acquisition parameters had different effect with a larger error for lower resolution (p=0.026), TR value (p=0.009) with a larger error for TR below 1500 ms; a flip angle value of 90 degrees was associated with higher error rates (p=0.006).

### Introspection results

When investigating in more details the misclassified cases (see Figure 5), it appeared that the characteristics of these gliomas reflected the texture properties of the classes they were misclassified into both with respect to the mutation status or the grading. For instance, similarly to what was observed between grade II and IV, the cases wrongly classified as grade IV instead of grade II had a significantly lower compactness, with higher mean and kurtosis and lower skewness as well as lower correlation and higher entropy among others. Numerical mean of z-score differences for each of the possible error cases are reported in Supplementary table 4.

**Figure 5:**
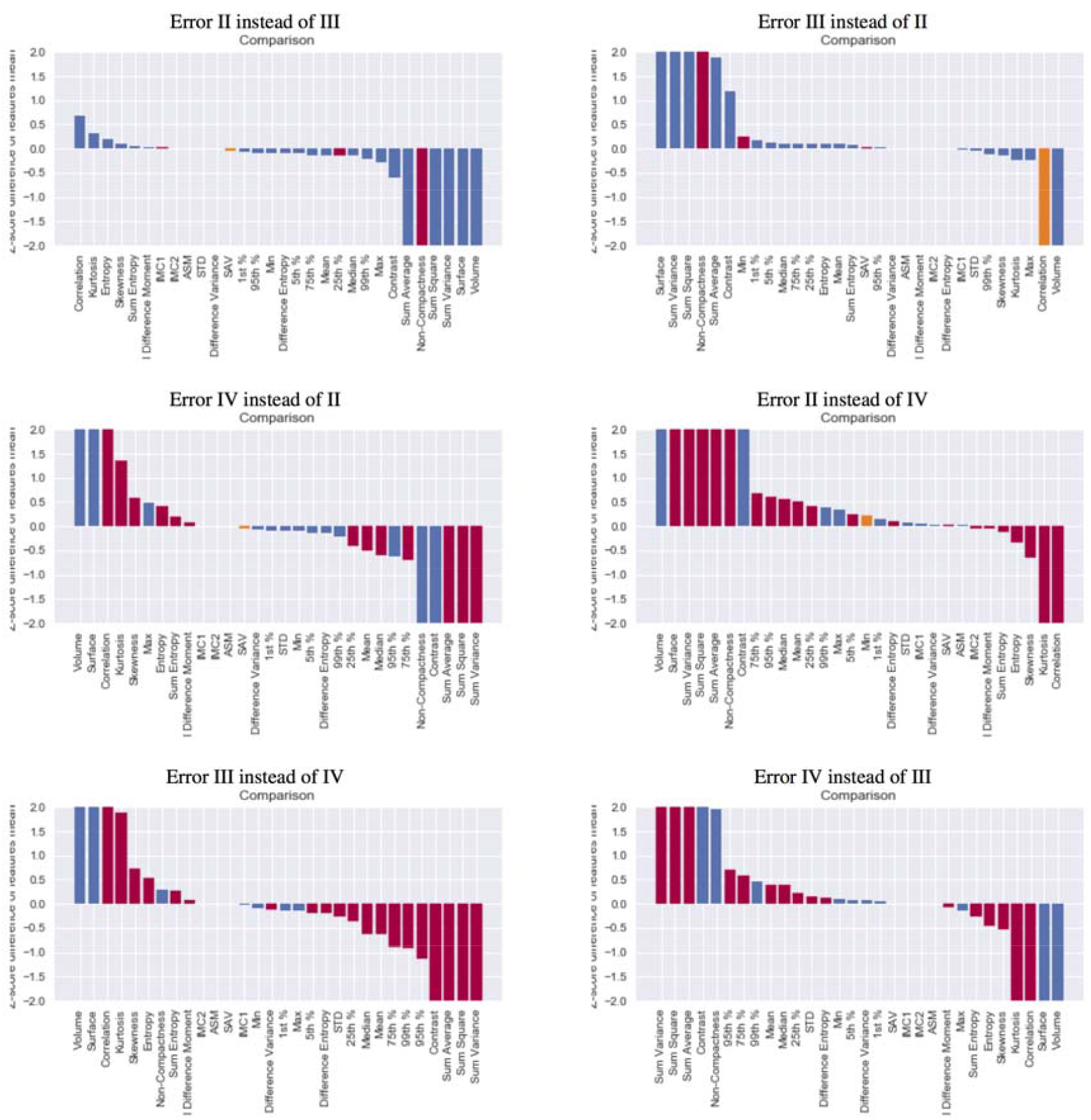
Comparison of features Z-Scores between erroneously classified and rightly classified elements for each possible error type. The bar length represents the Mean Z-Score difference between error cases and true cases. Bars in which difference was significant (Wilcoxon 2 sample test P<0.05) are coloured in red while others are in blue.

## Discussion

Our results suggest that the application of machine learning strategies to DSC-MRI extracted features, can successfully classify gliomas from multi-center patients pool by IDH mutation status and WHO grades II, III and IV. Applying a random-forest algorithm, we accurately predicted IDH mutation status in 72% of the cases with an overall specificity of 77% and sensitivity of 65%. On average, the algorithm classified 53% of the gliomas correctly into three histological grades II-IV, and with 87% the classification with an error distance of less than 1.

The novelty of our work is that we show that even simple shape features can help in the distinction between grade II and higher grades. The measure of non-compactness was significantly higher in WHO grade III or grade IV compared to grade II, reflecting possibly higher infiltrative features in these tumours. Interestingly, this shape feature was not found to be significantly different between WHO grade IV and grade III groups. It must be noted that in the existing literature, histogram features have generally been favoured against shape descriptors^36–38^. Falk et al. have found that 90^th^ percentile rCBV values were significantly lower in grade II gliomas than grade III (AUC=0.77, p=0.04), but they have also found the standard deviation of rCBF was the best discriminative feature with the highest AUC in the same setting (AUC=0.80, p=0.02). Our results do not confirm these findings, with histogram features only relevant markers of grade difference between grades IV and II or IV and III. Therefore, we suggest that shape descriptors, such as non-compactness and surface to volume ratio, have to be considered in the mapping of the composition of gliomas as they correlate well with histopathological findings. Perfusion texture features measuring the degree of intensity heterogeneity (such as the correlation or the sum of variance) appeared to be significantly different across the IDH mutation groups. Similar large number of markers of tissue heterogeneity were found to be different across a paired comparison of grades IV vs III and grades IV vs II. These observations are biologically relevant as they reflect heterogeneous microenvironment in high-grade gliomas, which is also consistent with diffusion studies^14,39^. It can be postulated that texture features reflect the continuum of appearance evolution between grades but the results highlight the challenge of strictly separating grades III from the two extremes of the spectrum (grade II and grade IV).

The prediction rate in our study performed better (65% of specificity, 79% sensitivity) than the one reported by Kickingereder et al.^8^ in a single-centre trial despite the overall heterogeneous nature of our large cohort in terms of molecular subgroups and DSC acquisition protocols. Similar machine-assisted techniques for tumour grade prediction demonstrated accuracy between 73% and 85%^40–43^. While our algorithm performance appears slightly inferior to the aforementioned results, we note that in all these studies patients are recruited from one centre, while we considered patients data from six centres. With multi-center data we expect higher variability and heterogeneity and hence anticipated lower accuracy. Using a different simulation approach might however improve accuracy. Citak-Er et al.^43^ have proposed the use of a linear kernel support vector machine (SVM) with ten-fold cross-validation based on features extracted from conventional and advanced MRI data (DTI, DWI, DSC perfusion and MR-Spectroscopy), and reported 73.3% of overall sensitivity from their multiparametric MR imaging. For the purpose of our study we chose random-forest algorithm as an established method for three classes or more classification. Future work would probably involve combining radiomics features from additional advanced MRI sequences so as to better identify required sequences for an optimised performance. With the recent rise of deep-learning techniques that do not rely on hand-crafted features, further exploration on this end would be required notably to assess the behaviour of such solutions in the identification of borderline cases, the associated uncertainty calibration as well as the assessment of robustness to acquisition changes.

Our error analysis was performed to better characterise the existing continuum between ordinal categories such as glioma grades. Analysis of the relationship between misclassifications and used features was strongly in agreement with previously well-known MRI shortcomings in tumour staging. For instance, some tumours incorrectly classified as WHO grade IV instead as grade III had significantly higher mean intensity in rCBV images. This highlights the inadequacy of first-order statistics to establish clear cut-off values regardless the sophistication of the DSC technique. This paradigm exemplifies the biological interpretability of machine-learning based radiomics compared to non-feature based methods and illustrates how it can be used to re-evaluate categorical classifications and improve characterisation and understanding of the pathological continuum in glioma grades, in particular with emerging novel, validated molecular biomarkers, such as CDKN2A/B deletion in IDH-mutant astrocytomas, which have been suggested to form a new clinical risk group^3^. Such use of radiomics techniques parallels the findings of new biological definition of GBM subgroups described in the work from Rathore et al^44^.

Overall, this proof-of-concept study aimed to assess the diagnostic value of the DSC-MRI radiomics in treatment-naïve gliomas using multi-centre study design without *a priori* consensus on the acquisition parameters and settings (incl. MR field strength, sequence type, resolution, acquisition parameters). Different suggestions for sequence optimisation in DSC-PWI, especially across TE and FA, are available in the literature^30^. The large protocol variability observed in our cohort allowed us to investigate the effect of these key characteristics. Acquisition metrics were dichotomized based on existing literature^30,45^, and subsequently analysed prediction results across defined groups. Higher acquisition resolution was significantly associated with lower error rates. Error in grade prediction was more sensitive to acquisition parameters than the distinction between IDH wild-type and mutant. Additionally, lower field strength associated with lower variability in scanning protocols led to lower absolute errors (p= 0.02 and p=0.01 for IDH and WHO grade respectively). Higher sensitivity and specificity rates with lower errors were achieved in the estimation of IDH mutation (82%, 70%, 0.26 at 1.5T, and 72%, 60%, 0.37 at 3T, respectively), and better WHO grade predictions, exact or with a distance, at 1.5T systems when compared to 3T. In fact, while higher spatial resolution and signal-to-noise ratio from higher magnetic fields may turn into an advantage in DSC imaging, which eventually leads decrease in the dose of contrast material and acquisitions time, protocol variability may increase image appearance heterogeneity. This result cannot be generalised in favour of the 1.5T field strength scanners as this would need a controlled randomised study but it stresses the need for imaging acquisition guidelines to harness the potential of technological advances, such as higher MRI fields. Last but not least, partial volume effect caused by surrounding tissues in the point of arterial input function may also result in erroneous estimation of rCBV.

The main strength of this work lies in both the multi-centre design and, though limited for optimised learning approaches, still the largest cohort in the literature to utilise machine-learning based radiomics as a predictive diagnostic tool. Since the analyses were purposefully performed across perfusion data only, further studies are needed to assess a combined impact of a multimodal approach (e.g. DWI, T1-weighted MR perfusion, MR spectroscopy, PET) in the imaging phenotyping in gliomas, though this carries the risk of introducing further considerable variability. The small number of patients in some groups did not allow to form integrated molecular subgroups, akin to previous work and undertake multi-class optimised learning classification according to the integrated histomolecular WHO classification scheme. Further data is continuously being gathered from the existing and new participating centres to extend the work in this direction.

In conclusion, the findings underscore the satisfactory diagnostic contribution of DSC-MRI extracted higher-order features combined with machine-learning in the automated classification of grading and IDH mutation status of gliomas mitigating the high imaging heterogeneity. The promising results obtained through the proposed random forest radiomics-based framework, are an additional step towards the clinical translation of machine learning tools as diagnostic aid aiming at facilitating the implementation of tailored treatment based on precision imaging

## Data Availability

Data and the machine learning algorithms used in this study are available at request from the authors.

## Author’s contributions

CHS, JPG, ES, MJC and SB significantly contributed to designing the study and drafted the paper. VKK, GS, FBP, CG, KSP, JA, MVS, MN, ARC, AA, SG, AK, NA, GMC, VR, LU, AE, EC,EG,EK, DR,JG and TB supplied the data for this study and contributed to improving the manuscript. All co-authors read and approved the submitted version of the manuscript.

## Data availability

Data and the machine learning algorithms used in this study are available at request from the authors subject to bilateral agreements.

## References

1. Yang M, Soga T, Oncometabolites PPJ. linking altered metabolism with cancer. J Clin Invest. 2013;123(9):3652–3658.

2. Louis DN, Perry A, Reifenberger G, von Deimling A, Figarella-Branger D, Cavenee WK and others. The 2016 World Health Organization Classification of Tumors of the Central Nervous System: a summary. Acta Neuropathol. 2016;131(6):803–820.

3. Shirahata M, Ono T, Stichel D, et al. Novel, improved grading system(s) for IDH-mutant astrocytic gliomas. Acta Neuropathol. 2018;136(1):153–166.

4. Chamberlain MC, Murovic JA, Levin VA. Absence of contrast enhancement on CT brain scans of patients with supratentorial malignant gliomas. Neurology. 1988;38(9):1371.

5. Scott JN, Pma B, Sevick RJ, Rewcastle NB, PA. F. How Often Are Nonenhancing Supratentorial Gliomas Malignant? A Population Study. Neurology; 2002.

6. Santarosa C, Castellano A, Conte GM, et al. Dynamic Contrast-Enhanced and Dynamic Susceptibility Contrast Perfusion MR Imaging for Glioma Grading: Preliminary Comparison of Vessel Compartment and Permeability Parameters Using Hotspot and Histogram Analysis. European Journal of Radiology; 2016.

7. Anzalone N, Castellano A, Cadioli M, et al. Brain Gliomas: Multicenter Standardized Assessment of Dynamic Contrast-enhanced and Dynamic Susceptibility Contrast MR Images. Radiology. 2018;1703:62-.

8. Kickingereder P, Bonekamp D, Nowosielski M, et al. Radiogenomics of Glioblastoma: Machine Learning-based Classification of Molecular Characteristics by Using Multiparametric and Multiregional MR Imaging Features. Radiology. 2016;281(3):907–918.

9. Hu LS, Kelm Z, Korfiatis P, et al. Impact of Software Modeling on the Accuracy of Perfusion MRI in Glioma. AJNR Am J Neuroradiol. 2015;36(12):2242–2249.

10. Kelm ZS, Korfiatis PD, Lingineni RK, et al. Variability and accuracy of different software packages for dynamic susceptibility contrast magnetic resonance imaging for distinguishing glioblastoma progression from pseudoprogression. J Med Imaging. 2015;2:2.

11. Conte GM, Castellano A, Altabella L, et al. Reproducibility of dynamic contrast-enhanced MRI and dynamic susceptibility contrast MRI in the study of brain gliomas: a comparison of data obtained using different commercial software. Radiol Med. 2017;122(4):294–302.

12. Welker K, Boxerman J, Kalnin A, et al. ASFNR recommendations for clinical performance of MR dynamic susceptibility contrast perfusion imaging of the brain. AJNR Am J Neuroradiol. 2015;36(6):E41–51.

13. Brynolfsson P, Nilsson D, Henriksson R, et al. ADC texture-An imaging biomarker for high-grade glioma? Med Phys. 2014;41(10):101903. doi:10.1118/1.4894812

14. Bisdas S, Shen H, Thust S, et al. Texture analysis- and support vector machine-assisted diffusional kurtosis imaging may allow in vivo gliomas grading and IDH-mutation status prediction: a preliminary study. Sci Rep. 2018;8:1.

15. Chaddad A, Zinn PO, Colen RR. Quantitative texture analysis for Glioblastoma phenotypes discrimination. In: 2014 International Conference on Control, Decision and Information Technologies (CoDIT). IEEE; 2014:605–608. doi:10.1109/CoDIT.2014.6996964

16. Yang D, Rao G, Martinez J, Veeraraghavan A, Rao A. Evaluation of tumor-derived MRI-texture features for discrimination of molecular subtypes and prediction of 12-month survival status in glioblastoma. Med Phys. 2015;42(11):6725–6735. doi:10.1118/1.4934373

17. McGarry SD, Hurrell SL, Kaczmarowski AL, et al. Magnetic Resonance Imaging-Based Radiomic Profiles Predict Patient Prognosis in Newly Diagnosed Glioblastoma Before Therapy. Tomography. 2016;2(3):223. doi:10.18383/J.TOM.2016.00250

18. Kickingereder P, Go tz M, Muschelli J, et al. Large-scale Radiomic Profiling of Recurrent Glioblastoma Identifies an Imaging Predictor for Stratifying Anti-Angiogenic Treatment Response. Clin Cancer Res. 2016;22(23):5765–5771. doi:10.1158/1078-0432.CCR-16-0702

19. Grossmann P, Narayan V, Chang K, et al. Quantitative imaging biomarkers for risk stratification of patients with recurrent glioblastoma treated with bevacizumab. Neuro Oncol. 2017;19(12):1688–1697. doi:10.1093/neuonc/nox092

20. Ertosun MG, Rubin DL. Automated Grading of Gliomas using Deep Learning in Digital Pathology Images: A modular approach with ensemble of convolutional neural networks. AMIA. Annu Symp proceedings AMIA Symp. 2015;2015:1899–1908.

21. Chang P, Grinband J, Weinberg BD, et al. Deep-Learning Convolutional Neural Networks Accurately Classify Genetic Mutations in Gliomas. Am J Neuroradiol. 2018;39(7):1201–1207. doi:10.3174/AJNR.A5667

22. Lu CF, Hsu FT, Hsieh KL, et al. Machine Learning-Based Radiomics for Molecular Subtyping of Gliomas. Clin Cancer Res. 2018;24(18):4429–4436.

23. Hartmann C, Meyer J, Balss J, et al. Type and frequency of IDH1 and IDH2 mutations are related to astrocytic and oligodendroglial differentiation and age: a study of 1,010 diffuse gliomas. Acta Neuropathol. 2009;118(4):469–474.

24. Louis DN, Perry A, Burger P, Ellison DW, Reifenberger G, von Deimling A and others. International Society Of Neuropathology--Haarlem consensus guidelines for nervous system tumor classification and grading. Brain Pathol. 2014;24(5):429–435.

25. Balss J, Meyer J, Mueller W, Korshunov A, Hartmann C, von Deimling A. Analysis of the IDH1 codon 132 mutation in brain tumors. Acta Neuropathol. 2008;116(6):597–602.

26. Brandner S, von Deimling A. Diagnostic. prognostic and predictive relevance of molecular markers in gliomas. Neuropathol Appl Neurobiol. 2015;41(6):694–720.

27. Jaunmuktane Z, Capper D, Dtw J, et al. Methylation array profiling of adult brain tumours: diagnostic outcomes in a large, single centre. Acta Neuropathol Commun. 2019;7:1.

28. Yushkevich PA, Piven J, Hazlett HC, et al. User-guided 3D active contour segmentation of anatomical structures: Significantly improved efficiency and reliability. Neuroimage. 2006;31(3):1116–1128.

29. Mouridsen K, Friston K, Hjort N, Gyldensted L, Østergaard L, Kiebel S. Bayesian estimation of cerebral perfusion using a physiological model of microvasculature. Neuroimage. 2006;33(2):570–579. doi:10.1016/j.neuroimage.2006.06.015

30. Boxerman JL, Paulson ES, Prah MA, KM. S. The effect of pulse sequence parameters and contrast agent dose on percentage signal recovery in DSC-MRI: implications for clinical applications. AJNR Am J Neuroradiol. 2013;34(7):1364–1369.

31. Boxerman JL, Schmainda KM, RM. W. Relative cerebral blood volume maps corrected for contrast agent extravasation significantly correlate with glioma tumor grade, whereas uncorrected maps do not. AJNR Am J Neuroradiol. 2006;34(7):1364–1369.

32. Modat M, Cash DM, Daga P, Winston GP, Duncan JS, Ourselin S. Global image registration using a symmetric block-matching approach. J Med Imaging. 2014;1(2).

33. Cardoso MJ, Modat M, Wolz R, et al. Geodesic Information Flows: Spatially-Variant Graphs and Their Application to Segmentation and Fusion. IEEE Trans Med Imaging. 2015;34(9):1976–1988. doi:10.1109/TMI.2015.2418298

34. Haralick RM, Shanmugam K, Dinstein I. Textural features for image Classification. IEEE Trans Syst Man Cybern Part B. 1973;SMC-3(6):610–621.

35. L. B. Statistical modeling: The two cultures. Stat Sci. 2001;16(3):199–215.

36. Catalaa I, Henry R, Dillon WP, et al. Perfusion, diffusion and spectroscopy values in newly diagnosed cerebral gliomas. NMR Biomed. 2006;19(4):463–475.

37. Kim H, Choi SH, Kim JH, et al. Gliomas: application of cumulative histogram analysis of normalized cerebral blood volume on 3 T MRI to tumor grading. PLoS One. 2013;8:5.

38. Falk A, Fahlstrom M, Rostrup E, et al. Discrimination between glioma grades II and III in suspected low-grade gliomas using dynamic contrast-enhanced and dynamic susceptibility contrast perfusion MR imaging: a histogram analysis approach. Neuroradiology. 2014;56(12):1031–1038.

39. Hempel JM, Schittenhelm J, Bisdas S, et al. In vivo assessment of tumor heterogeneity in WHO 2016 glioma grades using diffusion kurtosis imaging: Diagnostic performance and improvement of feasibility in routine clinical practice. J Neuroradiol. 2018;45(1):32–40.

40. Zöllner FG, Emblem KE, Lr S. Support vector machines in DSC-based glioma imaging: Suggestions for optimal characterization. Magn Reson Med. 2010;64:1230–1236.

41. Emblem KE, Due-Tonnessen P, Hald JK, Bjornerud A, Pinho MC, Scheie D and others. Machine learning in preoperative glioma MRI: survival associations by perfusion-based support vector machine outperforms traditional MRI. J Magn Reson Imaging. 2014;40(1):47–54.

42. Emblem KE, Pinho MC, Zollner FG, Due-Tonnessen P, Hald, J. K. and Schad LR and others. A generic support vector machine model for preoperative glioma survival associations. Radiology. 2015;275(1):228–234.

43. Citak-Er F, Firat Z, Kovanlikaya I, Ture U, Ozturk-Isik E. Machine-learning in grading of gliomas based on multi-parametric magnetic resonance imaging at 3T. Comput Biol Med. 2018;99:154–160.

44. Rathore S, Akbari H, Rozycki M, et al. Radiomic MRI signature reveals three distinct subtypes of glioblastoma with different clinical and molecular characteristics, offering prognostic value beyond IDH1. Sci Rep. 2018;8(1):5087. doi:10.1038/s41598-018-22739-2

45. Boxerman JL, Rosen BR, RM. W. Signal-to-noise analysis of cerebral blood volume maps from dynamic NMR imaging studies. J Magn Reson Imaging. 1997;7(3):528–537.

